# Mortality among Adults Ages 25-44 in the United States During the COVID-19 Pandemic

**DOI:** 10.1101/2020.10.21.20217174

**Authors:** Jeremy Samuel Faust, Harlan M. Krumholz, Katherine L. Dickerson, Zhenqiu Lin, Cleavon Gilman, Rochelle P. Walensky

## Abstract

**Introduction:** Coronavirus disease-19 (COVID-19) has caused a marked increase in all-cause deaths in the United States, mostly among adults aged 65 and older. Because younger adults have far lower infection fatality rates, less attention has been focused on the mortality burden of COVID-19 in this demographic.

**Methods:** We performed an observational cohort study using public data from the National Center for Health Statistics at the United States Centers for Disease Control and Prevention, and CDC Wonder. We analyzed all-cause mortality among adults ages 25-44 during the COVID-19 pandemic in the United States. Further, we compared COVID-19-related deaths in this age group during the pandemic period to all drug overdose deaths and opioid-specific overdose deaths in each of the ten Health and Human Services (HHS) regions during the corresponding period of 2018, the most recent year for which data are available.

**Results:** As of September 6, 2020, 74,027 all-cause deaths occurred among persons ages 25-44 years during the period from March 1st to July 31st, 2020, 14,155 more than during the same period of 2019, a 23% relative increase (incident rate ratio 1.23; 95% CI 1.21–1.24), with a peak of 30% occurring in May (IRR 1.30; 95% CI 1.27-1.33). In HHS Region 2 (New York, New Jersey), HHS Region 6 (Arkansas, Louisiana, New Mexico, Oklahoma, Texas), and HHS Region 9 (Arizona, California, Hawaii, Nevada), COVID-19 deaths exceeded 2018 unintentional opioid overdose deaths during at least one month. Combined, 2,450 COVID-19 deaths were recorded in these three regions during the pandemic period, compared to 2,445 opioid deaths during the same period of 2018.

**Meaning:** We find that COVID-19 has likely become the leading cause of death—surpassing unintentional overdoses—among young adults aged 25-44 in some areas of the United States during substantial COVID-19 outbreaks.

**Note:** *The data presented here have since been updated. As a result, an additional 1,902 all-cause deaths occurring among US adults ages 25-44 during the period of interest are not accounted for in this manuscript*.

## Introduction

COVID-19 has caused a marked increase in all-cause deaths in the United States, mostly among adults aged ≥65.^1,2^ Because younger adults have far lower case fatality rates, little attention has been focused on the mortality burden of COVID-19 in this demographic.

Accordingly, we sought to identify changes in all-cause mortality (excess deaths) and to determine years of lost life (YLL) among US adults aged 25-44. As the leading cause of death in this demographic prior to COVID-19 was drug overdoses, we compared the monthly incidence rates of COVID-19-specific deaths during the pandemic to unintentional overdose deaths (all drug and opioid-specific) during the corresponding period in 2018 in each of the ten Health and Human Services (HHS) regions.

## Methods

We performed an observational study by assembling the most recent publicly available data for all-cause mortality (2019–2020), unintentional drug overdose deaths (ICD10 X41-X44, Y11-Y15, 2018), unintentional opioid-specific deaths (*ibid* and T40.0-6, 2018), and COVID-19 deaths (ICD10 U071).^3,4^ We calculated death incident rates per 100,000 person-month with 95% confidence intervals (SAS 9.4, SAS Institute, Carey, NC).

We computed YLL due to COVID-19 and for all-cause excess mortality occurring from March– July 2020 to assess undetected burdens resulting from COVID-19 undercounting.^5^ Excess mortality was defined as deaths in 2020 minus 2019 deaths during the corresponding period. Incident rate ratios were also calculated.

This study was not subject to institutional review approval because it used publicly available data. STROBE reporting guidelines were followed.

## Results

From March–July 2020, 74,027 all-cause deaths occurred among persons ages 25-44 years, 14,155 more than during the same period of 2019, a 23% relative increase (incident rate ratio 1.23; 95% CI 1.21–1.24), with a peak of 30% occurring in May (IRR 1.30; 95% CI 1.27-1.33). In HHS Regions 2 (New York, New Jersey), HHS Region 6 (Arkansas, Louisiana, New Mexico, Oklahoma, Texas), and HHS Region 9 (Arizona, California, Hawaii, Nevada), COVID-19 deaths exceeded 2018 unintentional opioid overdose deaths during at least one month (Figure). Combined, 2,450 COVID-19 deaths were recorded in these three regions during the pandemic period, compared to 2,445 opioid deaths during the same period of 2018.

**Figure.**
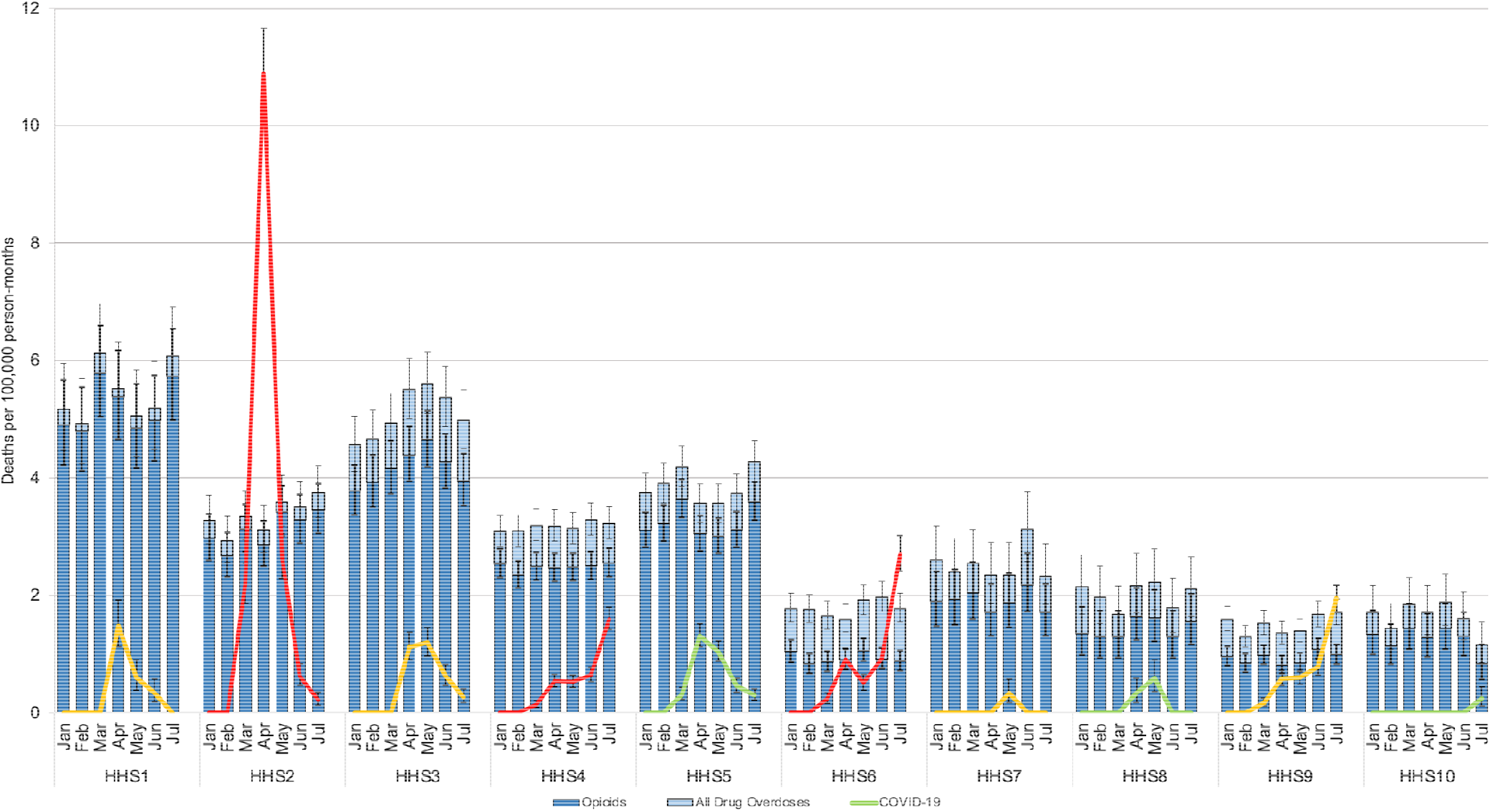
Unintentional overdose deaths, opioid deaths, and COVID-19 deaths among persons aged 25-44, per 100,000 person-month (vertical axis) by month (horizontal axis), stratified by HHS region. *Bars:* Unintentional drug overdose deaths (ICD10 X41-X45, Y11-Y15), 2018 (light blue with 95% CI) and opioid-specific drug overdose deaths (ICD10 X41-X45, Y11-Y15 *and* T40.0-40.6), 2018 (dark blue with 95% CI). *Lines:* COVID-19 deaths (ICD10 U071), 2020; colors are stratified by regional COVID-19 case incidence per 1,000,000 residents through July 31, 2020 (green <1,050 cases per 100,000 residents; yellow 1,050-1,499 cases per 100,000 residents; red >1,500 cases per 100,000 residents). HHS Regions: *Region 1:* Connecticut, Maine, Massachusetts, New Hampshire, Rhode Island, and Vermont; *Region 2*: New Jersey and New York; *Region 3*: Delaware, District of Columbia, Maryland, Pennsylvania, Virginia, and West Virginia; *Region 4*: Alabama, Florida, Georgia, Kentucky, Mississippi, North Carolina, South Carolina, and Tennessee; *Region 5*: Illinois, Indiana, Michigan, Minnesota, Ohio, and Wisconsin; *Region 6*: Arkansas, Louisiana, New Mexico, Oklahoma, and Texas; *Region 7*: Iowa, Kansas, Missouri, and Nebraska; *Region 8*: Colorado, Montana, North Dakota, South Dakota, Utah, and Wyoming; *Region 9*: Arizona, California, Hawaii, and Nevada; *Region 10*: Alaska, Idaho, Oregon, and Washington.

In March-July 2018, opioid overdoses caused 10,347 deaths nationwide, resulting in 472,608 YLL among adults 25-44. During the similar period in 2020, 4,055 recorded COVID-19 deaths in adults 25-44 resulted in 175,631 YLL (Table). Further considering all of the 14,155 excess deaths in 2020, younger adults accounted for 627,872 YLL, exceeding YLL from overdose-related deaths (595,175 YLL) during the corresponding period of 2018 (Table). Among adults 25-44, 28.7% (4,055 of 14,155) of the all-cause excess deaths were attributed to COVID-19.

**Table.** Years of life lost (YLL) due to opioid-specific overdose deaths, all overdose deaths, all-cause excess mortality, and COVID-19 in adults ages 25-44, March–July, 2020.

|  | Median life<br>expectation | Opioid<br>deaths<br>(2018) | YLL | All<br>overdose<br>deaths | YLL | Covid-19<br>deaths<br>(2020) | YLL | Excess deaths<br>(2020 versus<br>2019) | YLL |
| --- | --- | --- | --- | --- | --- | --- | --- | --- | --- |
| **Ages 25-44** |  |  |  |  |  |  |  |  |  |
| ages 25-34 | 50.0 | 5,536 | 276,800 | 6,746 | 337,300 | 1,139 | 56,950 | 5,566 | 278,300 |
| ages 35-44 | 40.7 | 4,811 | 195,808 | 6,336 | 257,875 | 2,916 | 118,681 | 8,589 | 349,572 |
| **total** |  | **10,347** | **472,608** | **13,082** | **595,175** | **4,055** | **175,631** | **14,155** | **627,872** |

Source: National Center for Health Statistics.<sup>3</sup>
Note: We used the most recently available public data; excess deaths (2020 and 2019), unintentional overdoses (2018).<sup>3,4</sup>

## Discussion

We find that COVID-19 has likely become the leading cause of death—surpassing unintentional overdoses—among young adults aged 25-44 in some areas of the United States with substantial COVID-19 outbreaks. In these regions, COVID-19 mortality also resembles that of the HIV/AIDS epidemic at its apex in the United States (1994-1995).^6^

We also found that a relatively small fraction of the all-cause excess deaths recorded during the pandemic have been attributed directly to COVID-19. This gap has not yet been explained, though a combination of inadequate testing and the continuation of pre-existing smaller increases in all-cause mortality recorded during January–February 2020 may explain a large fraction of this difference, implying that the mortality of COVID-19 has been substantially under-detected in the younger adult population.^1^

This study has limitations. The provisional data used represents lower-bound estimates due to reporting lags. While the magnitude of our findings may increase as COVID-19 spreads among younger populations, emerging treatments might improve fatality rates.

Together, these data indicate that the mortality of COVID-19 among adults ages 25-44 is sizeable and should not be discounted by individuals or policymakers.

## Data Availability

Dr. Faust had full access to all of the data in the study and takes responsibility for the integrity of the data and the accuracy of the data analysis.

## Data Statement and Author Contributions

*Concept and design:* Faust, Gilman, Krumholz, Walensky.

*Acquisition, analysis, or interpretation of data:* Faust, Dickerson.

*Drafting of the manuscript:* Faust, Walensky.

*Critical revision of the manuscript for important intellectual content:* All authors.

*Statistical analysis:* Faust, Lin, Krumholz, Walensky.

*Administrative, technical, or material support:* Dickerson.

*Supervision:* Faust, Krumholz, Walensky.

## Funding Statements

Faust: None.

Krumholz: Harlan Krumholz works under contract with the Centers for Medicare & Medicaid Services to support quality measurement programs; was a recipient of a research grant, through Yale, from Medtronic and the U.S. Food and Drug Administration to develop methods for post-market surveillance of medical devices; was a recipient of a research grant from Johnson & Johnson, through Yale University, to support clinical trial data sharing; was a recipient of a research agreement, through Yale University, from the Shenzhen Center for Health Information for work to advance intelligent disease prevention and health promotion; collaborates with the National Center for Cardiovascular Diseases in Beijing; receives payment from the Arnold & Porter Law Firm for work related to the Sanofi clopidogrel litigation, from the Martin Baughman Law Firm for work related to the Cook Celect IVC filter litigation and the Bard IVC filter litigation, and from the Siegfried and Jensen Law Firm for work related to Vioxx litigation; chairs a Cardiac Scientific Advisory Board for UnitedHealth; was a member of the IBM Watson Health Life Sciences Board; is a member of the Advisory Board for Element Science, the Healthcare Advisory Board for Facebook, and the Physician Advisory Board for Aetna; and is the co-founder of HugoHealth, a personal health information platform, and co-founder of Refactor Health, an enterprise healthcare AI-augmented data management company. He is also a consultant for FPrime.

Lin: Dr Lin reported working under contract with the Centers for Medicare & Medicaid Services.

Dickerson: None.

Gilman: .None.

Walensky: Dr. Walensky is the Stephen and Deborah Gorlin Research Scholar as Massachusetts General Hospital.

## References

1. Weinberger DM, Chen J, Cohen T, et al. Estimation of Excess Deaths Associated With the COVID-19 Pandemic in the United States, March to May 2020. JAMA Intern Med. Published online July 1, 2020. doi:10.1001/jamainternmed.2020.3391

2. Woolf SH, Chapman DA, Sabo RT, Weinberger DM, Hill L. Excess Deaths From COVID-19 and Other Causes, March-April 2020. JAMA. 2020;324(5):510–513. doi:10.1001/jama.2020.11787

3. COVID-19 Death Data and Resources - National Vital Statistics System. Published August 10, 2020. Accessed September 2, 2020. https://www.cdc.gov/nchs/covid19/covid-19-mortality-data-files.htm

4. Underlying Cause of Death, 1999-2018 Request Form. Accessed September 2, 2020. https://wonder.cdc.gov/controller/datarequest/D76

5. Arias E. United States Life Tables, 2016. National Vital Statistics Reports; vol 68 no 4 Hyattsville, MD: National Center for Health Statistics 2019.:66.

6. US Centers for Disease Control and Prevention. Update: Trends in AIDS Incidence, Deaths, and Prevalence -- United States, 1996. Accessed July 21, 2020. https://www.cdc.gov/mmwr/preview/mmwrhtml/00046531.htm#00002287.htm

